# Increased Mortality in Patients with Standard EEG Findings of “Diffuse Slowing”

**DOI:** 10.1101/19009621

**Authors:** Rob Wanzek, Nicholas Bormann, Yaseen Shabbir, Taku Saito, Thoru Yamada, Gen Shinozaki

## Abstract

**Background/Objectives:** We aim to confirm the association between the high risk score on bispectral electroencephalogram (BSEEG) and mortality by comparing outcomes for those with “diffuse slowing” and normal findings on standard EEG.

**Design:** This is a retrospective study conducted with patient chart data from March 2015 to March 2017.

**Setting:** Single center study at a tertiary care academic hospital in the Midwest region of the USA.

**Participants:** 1069 subjects aged 55 years and older who were on an inpatient floor or intensive care unit and received a standard 24-hour EEG.

**Measurements:** Primary outcome was all-cause mortality at 30-, 90-, 180-, and 365-days. Secondary outcomes were time-to-discharge, and discharge to home.

**Results:** Patients with “diffuse slowing” on standard EEG was significantly associated with 30-, 90-, 180-, and 365-day mortality (P < .001) compared to those with normal EEG findings when controlling for age, sex, and Carlson Comorbidity Index. Those with diffuse slowing also had a longer time to discharge (P < 0.001) and were less likely to discharge to home (P < 0.001) when controlling for the same factors. Findings were similar when limiting the study to only patients whose clinical status indicated “awake” at time of EEG, except for 30-day mortality.

**Conclusion:** Our findings show that a standard EEG finding of “diffuse slowing” for inpatients 55 year or older is associated with greater mortality. This study strengthens the importance of the association found between high BSEEG score and mortality.

## Introduction

Delirium is defined as an acute decline in attention and either disorganized thinking or altered level of consciousness with a fluctuating course.(1) Delirium can be considered to be acute brain failure(2) and is thought to be caused by many factors, including drugs, infections, central nervous system (CNS) insults,(3) sleep disturbance,(4) metabolic disturbances(5), and pain.(6) Advanced age and comorbidities predispose patients to developing delirium following any of the above insults.(7) (8) Delirium is very common in older patients, complicating about a third of hospital stays and often persists following discharge.(9, 10) Moreover, it is associated with higher morality and worse functional outcomes.(11) Thus, identification of delirious patients to initiate treatment for potentially reversible causes is vital to improve patient outcomes.

Delirium is typically identified by clinical assessment using the Confusion Assessment Method (CAM)(12) or similar questionnaire style instruments meant particularly for settings such as the intensive care unit (CAM-ICU).(13) However, it is frequently underdiagnosed in the hospital because of its often subtle and varied presentations; agitated, hyperactive delirium represents the minority of cases while mixed and hypoactive delirium are more common.(9) Reliable biomarkers of delirium are therefore desirable and needed for better patient care.

Electrophysiological brain signals are a well-known biomarker, with a specificity of 91% in one study of patients with dementia.(2) In that study, electroencephalogram (EEG) had a low sensitivity but another study with a more general population found that 96% (50/52) of those with encephalopathy have “background slowing” indicating that EEG signals read by neurology experts as “diffuse slowing” is a sensitive marker for encephalopathy.(14)

A finding of “diffuse slowing” on standard EEG is a characteristic feature of delirium and helpful in identifying delirious patients, including those with an underlying cognitive impairment such as dementia.^(2, 15)^ Thus, use of EEG could facilitate early identification of delirium. However, standard EEG is a burdensome procedure with many technical requirements.(16) Recently our group developed a screening tool for slow brain wave detection, a bi-spectral bedside EEG (BSEEG) and showed that high BSEEG scores were significantly associated with delirium.(17, 18) Further, our additional study using the BSEEG identified an association between high BSEEG score and mortality among elderly inpatients.(19) To confirm the association between slow brain wave and high mortality, this study aimed to test the association by investigating patient outcomes for those with “diffuse slowing” on standard EEG.

## Methods

### Study Oversight

This is a retrospective cohort study to determine the association between EEG findings of “diffuse slowing” and patient outcomes such as mortality. This study was approved by the University of Iowa Institutional Review Board.

### Patient Population

For the purpose of this study, we reviewed all patients’ records who received a standard 24-hour EEG as an inpatient at the University of Iowa Hospitals and Clinics (UIHC) from March 2015 to March 2017. The study team selected potential subjects based on age, including those 55 years or older in an effort to focus on an elderly population, knowing that this population is more susceptible to delirium and mortality.

### EEG Data Collection

1069 bedside 24-hr standard EEGs completed on inpatients age 55 years or older at UIHC between March 1^st^, 2015, and March 1^st^, 2017 were identified. The first day of EEG recording was used in our analysis and EEGs from subsequent days were not included. Subjects included patients from both ICU and medical floors. Age, sex, date of admission, date of EEG, date of discharge, mortality status, and date of death were recorded for the subject of each report with a follow-up period of one year. In addition, lists of subjects’ ICD 10 diagnoses were recorded using data from index admission and all previous admissions. To adjust for the effect of subjects’ comorbidities on mortality, Charlson Comorbidity Index (CCI) scores were calculated using subjects’ ICD10 codes.(20, 21) For each EEG performed, a corresponding report documented by neurologists specialized in electrophysiology was extracted. EEG reports included data on indication for study, clinical state, EEG recording including background, ictal and interictal discharges, EEG reactivity, clinical (video) events, description of findings, and neurologist impression. Based on the neurologist descriptions and impressions, reports were manually coded by the research team as having (a) a finding of diffuse slowing, diffuse delta and theta waves, background slowing, or similar terms, (b) a focal finding or lateralization, and (c) having ictal or seizure findings in fitting with standard terminology.(22)

At the time of EEG recording, the 1069 subjects included were on average 69.5 years old, median 68 years old, standard deviation 9.7. 53.5% of subjects were male. Average CCI score was 3.6, median 3, range 0 to 18. Brief reasons given for EEG requests included evaluation for seizure in 803 (75.1%) subjects, and some form of altered mental status for 285 (26.7%) subjects.

### Outcome Measures

Primary outcome was 30-, 90-, 180-, and 365-day mortality and was identified using chart review and obituary record. Secondary outcomes included time to discharge following EEG, and discharge disposition either to home or not to home.

### Statistical Analysis

Multivariate regression models were used to determine the association between findings and outcomes controlling for age, sex, and Charlson Comorbidity Index (CCI) score. In additional analysis, those with diffuse slowing were stratified by clinical status into two groups: those whose clinical status indicated “awake” and those whose clinical status did not indicate “awake.” Two-sided P-values of 0.05 or less were considered to indicate statistical significance. Statistical analysis was performed using RStudio software, version 1.2.1335.

## Results

### Group Comparisons

Categorizing subjects, 707 (66.1%) had diffuse slowing, 75 (7.0%) had ictal findings, i.e. seizures, 502 (47.0%) had some focal finding or lateralization of findings, and 119 (11.1%) had a normal EEG. There were 372 (34.8%) with diffuse slowing and no focal or seizure findings, 291 (27.2%) with diffuse slowing and some focal finding or lateralization but no seizure, and 145 (13.6%) with a focal finding but no diffuse slowing or seizure.

At 1 year, 15/119 (12.6%) with normal EEG had died, 41/145 (28.2%) of those with focal findings without diffuse slowing or ictal findings, 138/372 (37.1%) of those with diffuse slowing without focal or ictal findings, 122/291 (42.0%) of those with diffuse slowing, a focal finding or lateralization and no ictal finding, and 32/75 (42.7%) of those with ictal findings died. Mortality at earlier time intervals is visualized with a survival curve, Figure 1, which also displays the number at risk at intervals of 60 days.

**Figure 1.**
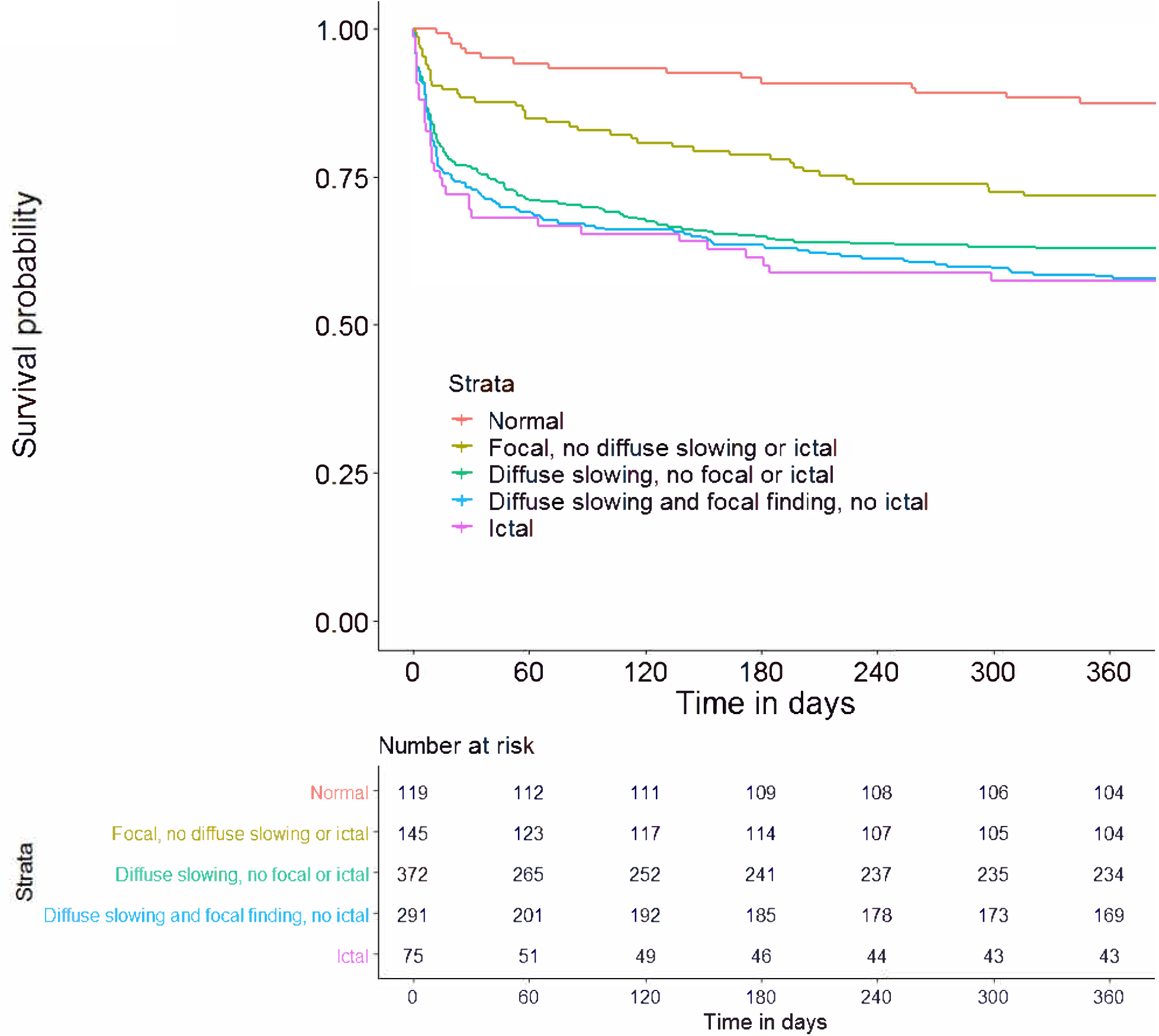

At 1 year, all groups with abnormal findings appear to do worse than those with normal findings (salmon-colored), with ictal and diffuse slowing apparently similar to one another (teal, blue, and violet), and worse than those with only focal findings (yellow). These differences are significant controlling for age, sex, and CCI except that the difference between the diffuse slowing nonfocal group is not significantly worse than the focal only group (P =0.148), as shown in Table 1.

**Table 1.** P values for group comparisons at 1 year.

|  | **DS+Focal** | **DS+nonfocal** | **Ictal** | **Focal** |
| --- | --- | --- | --- | --- |
| **DS+Focal** |  |  |  |  |
| **DS nonfocal** | 0.147 |  |  |  |
| **Ictal** | 0.706 | 0.138 |  |  |
| **Focal** | 0.00990 ** | 0.148 | 0.0250 * |  |
| **Normal** | 1.08e-05 *** | 0.000295 *** | 5.55e-05 *** | 0.0293 * |

P values for multivariate linear regression controlling for age, sex, and CCI.

### Diffuse Slowing Analysis

Because the two diffuse slowing groups were similar, we combined the two for further analysis of mortality and secondary outcomes. We stratified the combined diffuse slowing group by clinical status into two groups: those whose status included “awake” during EEG recording and those whose status did not. The survival curve shown in Figure 2 shows the difference from normals is retained in the awake group and shows a worse outcome among those with diffuse slowing who were not awake. Multivariate regressions controlling for age, sex, and CCI confirm that the difference between the awake group and normals is significant at 90 days and beyond. Table 2 shows results of statistical analysis for mortality outcomes as well as secondary outcomes, which were also significant. The other group is significantly different from both normals and from the awake group (P values listed in Table 2 are those for difference from the awake group).

**Table 2.** Primary and Secondary Outcomes for Diffuse Slowing groups.

|  | Normal (119) | Diffuse Slowing: Awake (289) | Diffuse Slowing: other status (374) |
| --- | --- | --- | --- |
| **30 day Mortality** | 5 (4.2%) | 35 (12.1%) (ns) | 130 (34.8%) *** |
| **90 day Mortality** | 8 (6.7%) | 59 (20.4%) * | 150 (40.1%) *** |
| **180 day Mortality** | 10 (8.4%) | 76 (26.3%) * | 161 (44.7%) *** |
| **1 Year Mortality** | 15 (12.6%) | 86 (29.8%) * | 174 (46.5%) *** |
| **Time to Discharge (days)** | Average: 4.6<br>Median: 2 | Average: 7.2<br>Median: 5<br>** | Average: 11.5<br>Median: 8<br>*** |
| **Discharge to home** | 92 (77.3%) | 100 (34.6%) *** | 63 (16.8%) *** |

The diffuse slowing groups are a combination of the focal and nonfocal groups from above and do not have ictal findings. Results of multivariate regression controlling for age, sex, and CCI comparing the awake group to normals and the other group to the awake group. P values:
\*<0.05, \*\*<0.01, \*\*\*<0.001. Not significant = (ns)

**Figure 2.**
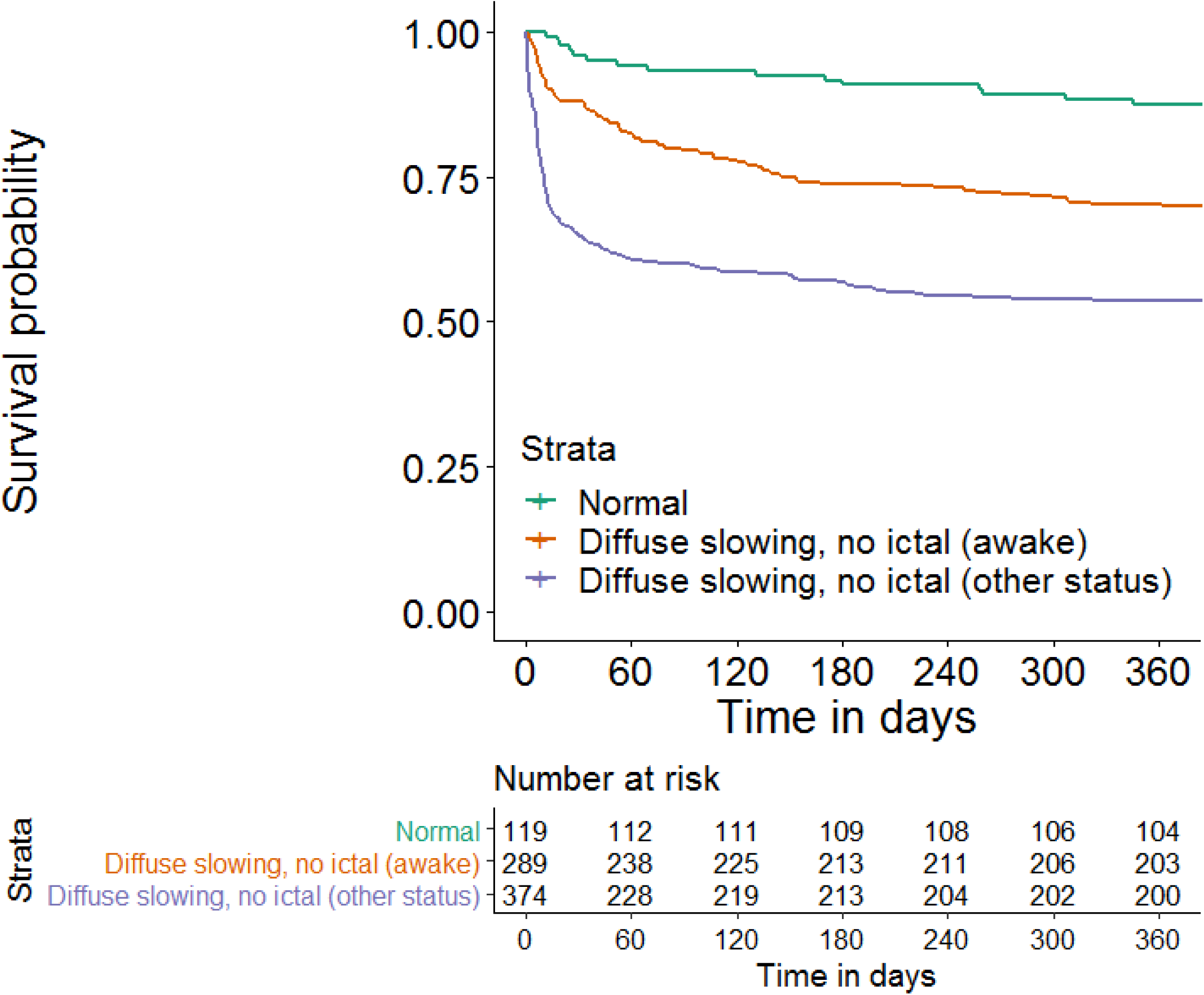

## Discussion

Continuous EEG (cEEG) monitoring is reportedly used in close to 1/100 mechanically ventilated patients based on a study from 2008(23) but likely is more common, with another study finding a 10-fold increase from 2004 to 2013.^(24)^ Continuous EEG offers much better detection of seizure than routine EEG,(25, 26) which is important because mortality is much higher in nonconvulsive seizures (32-51%), than in those without (13%).(27) While one study did not show better discharge Glasgow Coma Scale (GCS) score in those who received cEEG compared to those who did not, the study did note a difference in GCS at *admission*, which is consistent with an improvement related to EEG use although such a retrospective study does not allow for inference of causation.(28) Other studies do show differences in in-hospital mortality(23, 27) with use of cEEG, and a randomized control study investigating differences in 6-month mortality between use of routine and continuous EEG is underway.(29)

While investigation for seizure is often the stated indication for a continuous EEG study(30) “diffuse slowing” is a common finding and may be given less attention than needed. In our retrospective study of standard, 24-hour EEG reports in 1069 inpatients, we found that “diffuse slowing” on cEEG is significantly associated with mortality after an adjustment for age, sex, and CCI, confirming previous associations between diffuse slowing on EEG and mortality.(31, 32) In addition, we demonstrated the association persists even when limited to awake patients only. Our results also indicated that the mortality among the group with “diffuse slowing” is as bad as that among those with seizure and is worse than those with only focal findings. That the association becomes significant at 90 days and continues to 1 year in awake patients suggests that diffuse slowing is not necessarily an acute process which resolves but may be a marker of a more continual effect beyond its occurrence during hospitalization. Diffuse slowing may be important to screen for beyond its usual identification as an incidental finding in those evaluated for seizures. Our study therefore confirms the importance of previously mentioned association between bispectral EEG and mortality.(19)

Indications for EEG and frequency of outcomes were comparable to a large, three center study.(33) In this study 75.1% or orders were for underlying seizures while in that study 72.5%-70% of indications were for nonconvulsive seizures. There were also similar rates of seizures, with this study finding seizures at a rate of 7.0% using the first 24 hours of continuous EEG while that study found seizures at a rate of 12.9% over multiple day recordings, stating around 87.2% of seizures were found in the first day for a 24-hr rate of 11.2%. The difference may be due to physician ordering patterns.

There were several limitations to the study. The first of which is a feature of the finding of diffuse slowing itself. Anesthetic agents are very common and can cause diffuse slowing on EEG. However, we did not collect data on the use of anesthetic agents such as propofol or benzodiazepines or excluding subjects receiving anesthetic agents because doing so would likely eliminate many who had diffuse slowing for some other reason, or whose slowing was multiply-determined. Our study of awake patients mitigates that problem to some extent, as awake patients are less likely to have received sedatives, and we can be more confident that those in that group had a finding of diffuse slowing for some other reasons.

Other limitations relate to the generalizability of the study and to treatment applications. The study was performed at a single center. EEGs used in this study were not ordered at a standardized time or for a standardized indication, and results may have been affected by different ordering habits of treating physicians. This study did not include outpatients and the finding of diffuse slowing may not be associated with mortality in that population. This study did not determine the meaning of diffuse slowing in biological, biochemical, or anatomical terms. This study did not compare other potential factors influencing EEG findings such as specific medical/neurological conditions, medications, or interventions they received following EEG findings, although inclusion of CCI was meant to help control for comorbidity factors. CCI is limited in that it does not distinguish between conditions occurring before admission or during admission so it is unclear if or how they affected the use of EEG. We did not compare patients who didn’t receive EEGs and/or those who received only routine EEGs. This study does not demonstrate that identifying diffuse slowing or other abnormal findings leads to changes in management or outcomes.

## Conclusion

Our findings show that an EEG finding of “diffuse slowing” in the inpatient setting for patients 55 year or older is associated with greater mortality, in fact similar to mortality found in those with seizures. Our study suggests that the finding of diffuse slowing on EEG, which is a characteristic EEG feature for delirium, is an important clinical marker for predicting mortality and therefore bispectral EEG may become a useful screening tool for elderly patients if identification of diffuse slowing can lead to better outcomes.

## Data Availability

Data is available from author upon request.

## Declaration of Funding and Acknowledgements

Gen Shinozaki is a co-founder of Predelix Medical LLC. Other authors have no disclosures.

We certify that this work is novel of recent novel clinical research.

## Author Contributions

GS conceptualized and designed the study. RW and TY acquired the data. RW, NB, YS, and TS analyzed the data. RW drafted the initial manuscript, and GS, NB, YS, TS, and TY revised it. All authors have been listed who contributed significantly to the work and have approved the final version of the manuscript.

